# Global trends and vaccine-associated changes in meningitis burden, 2000–2023: a multi-country ecological panel study

**DOI:** 10.64898/2026.09.14.26363017

**Authors:** Khalid Mohammed Al-Dhayani

**Author notes:** **Corresponding Author:** Khalid Mohammed Al-Dhayani, Faculty of Medicine, Amran University, Amran City, Yemen.

## Abstract

**Background:** Meningitis remains a major cause of global morbidity and mortality. We aimed to quantify the association between vaccine introduction and meningitis-related disability-adjusted life year (DALY) rates globally and across high-burden regions from 2000 to 2023.

**Methods:** We conducted a longitudinal panel study of 188 countries using health metrics from the Global Burden of Disease study and WHO vaccine data. We estimated associations using two-way fixed effects regression models, adjusting for GDP and healthcare access. Analyses were stratified by baseline burden and included the African meningitis belt.

**Findings:** Globally, meningitis DALY rates declined by 4.97% per year (p<0.0001). Introduction of the Hib vaccine was associated with a 6.7% reduction in DALY rates, and PCV with a 4.8% reduction. In high-burden countries, both vaccines were associated with larger reductions (∼10%), while no significant effects were detected in low-burden settings. In the African meningitis belt, PCV introduction was associated with a substantial 16.8% reduction in DALY rates (p<0.0001). No significant global association was observed for meningococcal vaccines.

**Interpretation:** Global declines in meningitis burden are strongly associated with Hib and PCV scale-up, with the greatest benefits in high-burden settings. These findings support prioritized vaccine deployment in endemic regions to maximize public health impact. However, the presence of pre-treatment trends for Hib suggests these estimates represent vaccine-associated population-level changes rather than strictly causal effects.

## 1. Introduction

Meningitis remains a significant global public health concern, contributing to high morbidity and mortality, particularly among children under five years of age[1,2]. The burden of disease, often quantified in Disability-Adjusted Life Years (DALYs), is driven primarily by premature mortality (Years of Life Lost, YLL) and, to a lesser extent, long-term neurological sequelae (Years Lived with Disability, YLD)[3,4]. Despite substantial progress in reducing overall meningitis burden over the past two decades, incidence remains unevenly distributed, with low- and middle-income countries—particularly in sub-Saharan Africa—bearing the highest disease load[5,6]. Vaccination is recognized as the most effective preventive measure against bacterial meningitis. Haemophilus influenzae type b (Hib) vaccines, introduced in many countries since the early 2000s, have substantially reduced invasive Hib disease and related deaths[7]. Pneumococcal conjugate vaccine (PCV) and meningococcal (Men) vaccines (e.g., MenA, MenACWY) further contribute to controlling disease caused by Streptococcus pneumoniae and Neisseria meningitidis, respectively[8,9]. While Hib and PCV have been widely integrated into routine immunization schedules, Men vaccines in endemic regions such as the African meningitis belt are often deployed through mass campaigns, which can result in heterogeneous temporal coverage and impact[10,11].

Existing literature primarily reports descriptive trends in meningitis burden or vaccine coverage[3,4,6]. Few studies quantitatively link vaccine introduction to reductions in DALYs using longitudinal, multi-country panel data. Addressing this gap is critical for understanding both global and regional variations in vaccine effectiveness and for informing targeted public health strategies[12,13].

In this study, we integrate 24 years (2000–2023) of country-level meningitis DALYs with national vaccine introduction data for Hib, PCV, and Men vaccines. Using a Two-Way Fixed Effects econometric framework, we estimate the impact of these interventions on the global burden of meningitis and explore heterogeneity across high- and low-burden settings, with a focused analysis on the African meningitis belt.

## 2. Methods

### 2.1. Data Source Consolidation

This longitudinal study integrates two primary datasets covering the period 2000– 2023. Health metrics, including DALYs, YLL, and YLD, were extracted from the Institute for Health Metrics and Evaluation (IHME) GBD database[2,4]. Vaccine implementation data for Hib, PCV, and Men vaccines were obtained from the WHO Immunization Dashboard[14]. Data from over 190 countries and territories were merged into a longitudinal country-year panel dataset of meningitis burden and preventative interventions.

### 2.2. Outcome Definition and Data Processing

The primary outcome was the annual meningitis DALY rate per 100,000 population. Given the characteristic right-skewness of global health metrics, DALY, YLL, and YLD rates underwent natural log-transformation[3]. This transformation stabilizes variance, satisfies linearity assumptions for parametric modeling, and allows for the interpretation of regression coefficients as semi-elasticities. Records with zero values for health outcomes were excluded to avoid computational artifacts during transformation. The visual comparison of distributional properties, demonstrating the substantial reduction in skewness and kurtosis before and after transformation, is detailed in Supplementary Figure S1[13]. DALY, YLL, and YLD measures were extracted from the GBD 2021 Results Tool as age-standardised rates per 100,000 population for all sexes combined.

### 2.3. Geospatial Mapping and Relational Merging

Country names were standardized using ISO-3166-1 alpha-3 codes through programmatic mapping with the pycountry Python library[15]. To maintain sovereign-level analysis and reduce geographic misclassification, sub-national regions and supra-national aggregates were excluded, representing the largest reduction in the initial dataset (Figure 1). Sub-national regions and supra-national aggregates were excluded to maintain sovereign-level analysis, reducing potential geographic misclassification. This step ensured precise relational merging between health outcomes and vaccine introduction data.

**Figure 1.**
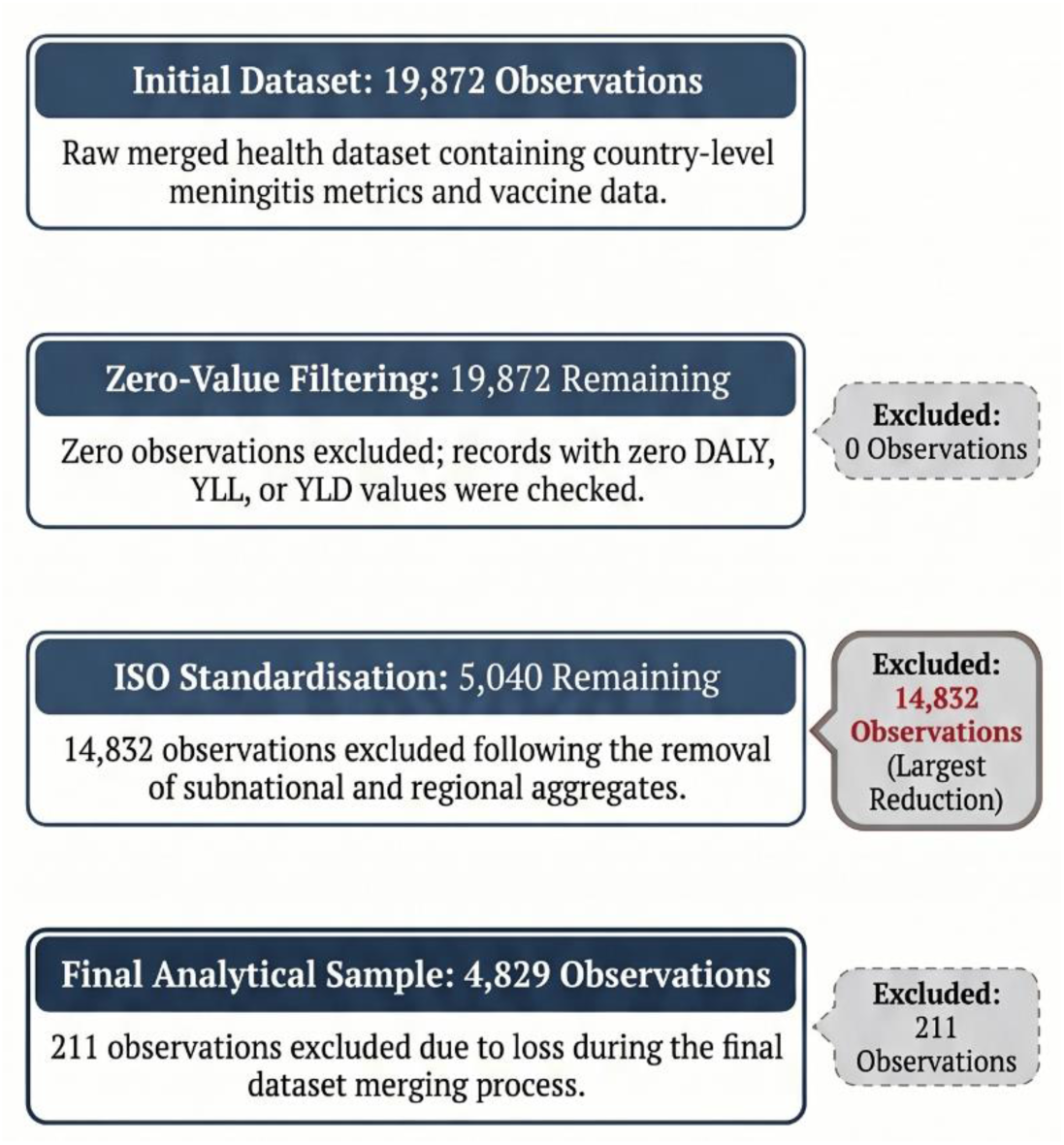
Data processing and sample attrition flowchart. The flowchart illustrates the systematic exclusion criteria used to derive the final analytical sample of 4,829 country-year observations. The initial dataset of 19,872 records was refined through quality control and geographic standardization. The largest reduction (n=14,832) resulted from the exclusion of subnational regions and supra-national aggregates to ensure analysis at the sovereign level only. A further 211 observations were lost during the final relational merging between health metrics and vaccine implementation data.

### 2.4. Econometric Specification

We employed a Two-Way Fixed Effects (TWFE) panel regression model to estimate the impact of vaccine introduction on log-transformed DALYs:

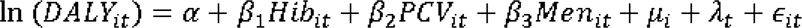

where *µ_i_* represents country-specific fixed effects, controlling for time-invariant heterogeneity, and *λ_t_* represents year fixed effects, accounting for global temporal shocks[12]. Models were estimated using the linearmodels.PanelOLS package in Python, with standard errors clustered at the country level to mitigate within-country serial correlation[16]. The parallel trends assumption was evaluated using event-study specifications examining pre-intervention coefficients relative to vaccine introduction timing. Because vaccine adoption occurred asynchronously across countries, estimates were interpreted cautiously in light of recent literature describing potential biases in conventional two-way fixed effects estimators under staggered treatment timing and heterogeneous treatment effects[17,21,22]. Event-study specifications additionally incorporated country-specific linear time trends to account for differential secular trajectories in meningitis burden across countries. This approach was used to reduce bias arising from heterogeneous pre-intervention trends in vaccine adoption timing.

### 2.5. Stratification and Subgroup Analysis

To assess heterogeneity, countries were stratified into High-Burden and Low-Burden cohorts based on median baseline DALY rates[5]. A targeted analysis focused on 15 countries within the African meningitis belt, a region historically characterized by recurrent meningococcal epidemics[6]. This stratification enables evaluation of how baseline disease intensity and geographic context modify vaccine effectiveness.

### 2.6. Statistical Considerations

Log-transformations were applied to reduce skewness and kurtosis in DALY, YLL, and YLD rates, ensuring OLS assumptions of normality and homoscedasticity were met[13]. Vaccine indicators (Hib_Post, PCV_Post, Men_Post) were coded as binary (0 = pre-introduction, 1 = post-introduction) to model semi-elasticities of disease reduction[12]. Sample attrition due to dataset merging and subnational exclusion is detailed in Supplementary Table S1. Model diagnostics included R-squared (within, between, overall), F-tests, and examination of clustered standard errors to validate inference reliability[16]. Additional sensitivity analyses incorporated time-varying country-level covariates, including log GDP per capita and Healthcare Access and Quality Index (HAQI) scores, to evaluate the robustness of estimated vaccine associations to changes in socioeconomic development and healthcare system capacity over time.

## 3. Results

### 3.1 Study population

The final analytical sample comprised 4,829 country-year observations from 188 countries and territories, following the exclusion of records lacking sovereign-level ISO-3 codes or those with incomplete relational merging (Figure 1). Baseline characteristics for these countries are summarized in Table 1.

**Table 1.** Baseline characteristics of included countries, 2000–2023.

| Characteristic | All countries<br>(n=188) | High-burden<br>(n=94) | Low-burden<br>(n=94) |
| --- | --- | --- | --- |
| **Meningitis burden**<br>**(rate per 100,000)** |  |  |  |
| Median DALY rate (IQR) | 79.50 (29.79–586.03) | 587.72 (156.39–1588.50) | 29.79 (18.09–50.75) |
| Median YLL rate (IQR) | 22473.11 (17441.30–36853.14) | 36461.54 (22927.76–55855.92) | 18089.18 (14486.02–22214.13) |
| Median YLD rate (IQR) | 10938.03 (9874.57–12754.77) | 10076.01 (9485.38–10795.06) | 12686.84 (11171.68–13831.62) |
| **Vaccine introduction**<br>**year, median (range)** |  |  |  |
| Hib vaccine | 2007 (2000–2019) | 2007 (2000–2019) | 2007 (2000–2014) |
| PCV | 2012 (2000–2023) | 2014 (2004–2023) | 2011 (2000–2023) |
| Men vaccine | 2012 (2000–2023) | 2016 (2003–2023) | 2010 (2000–2022) |
| Average population, millions (SD) | 16.46 (5.83) | 16.59 (5.86) | 16.33 (5.80) |
| GDP per capita, median (IQR) | 6716.10 (5656.94–7721.49) | 6744.54 (5622.95–7715.30) | 6687.47 (5689.88–7725.79) |
| HAQI score, median (SD) | 59.38 (11.90) | 59.29 (11.72) | 59.56 (12.07) |
| **WHO region, % (n)** |  |  |  |
| African Region | 16.5% (31) | 14.9% (14) | 18.1% (17) |
| Region of the Americas | 14.9% (28) | 13.8% (13) | 16.0% (15) |
| South-East Asia Region | 14.9% (28) | 13.8% (13) | 16.0% (15) |
| European Region | 21.8% (41) | 21.3% (20) | 22.3% (21) |
| Eastern Mediterranean Region | 16.5% (31) | 18.1% (17) | 14.9% (14) |
| Western Pacific Region | 15.4% (29) | 18.1% (17) | 12.8% (12) |

**Abbreviations:** DALY, disability-adjusted life years; YLL, years of life lost; YLD, years lived with disability; HAQI, Healthcare Access and Quality Index; IQR, interquartile range; SD, standard deviation. High- and low-burden groups were defined according to the median baseline DALY rate.

**Table 2.** Association between vaccine introduction and meningitis DALY rates stratified by baseline disease burden.

| Variable | High-burden countries | Low-burden countries |
| --- | --- | --- |
| Hib vaccine (post) | −0.0962*** (−3.26) | 0.0176 (0.42) |
| PCV (post) | −0.0949*** (−2.79) | −0.0049 (−0.25) |
| Men vaccine (post) | 0.0541 (1.00) | −0.0276 (−0.84) |
| Observations | 2415 | 2414 |
| Within R<sup>2</sup> | 0.266 | 0.018 |
| Fixed effects | Country, year | Country, year |

**Data are $\beta$ coefficients (t statistics).** \*\*\*p<0.001, \*\*p<0.01, \*p<0.05. Stratified analysis of vaccine impact on meningitis DALY rates by baseline disease burden. Countries were classified as high or low burden based on the median baseline DALY rate. Models include country and year fixed effects with clustered standard errors.

### 3.2. Temporal Trends and Structural Drivers of Meningitis Burden

Between 2000 and 2023, meningitis-related DALY rates declined consistently across countries (Figure 2). In TWFE models controlling for country and year effects (Table 3, Model 2), each additional year was associated with a 4.97% reduction in DALY rates (β = −0.0497, p < 0.001). Globally, the meningitis burden remained predominantly driven by premature mortality (YLL), with disability-related burden (YLD) comprising a smaller proportion of total DALYs throughout the study period.

**Figure 2.**
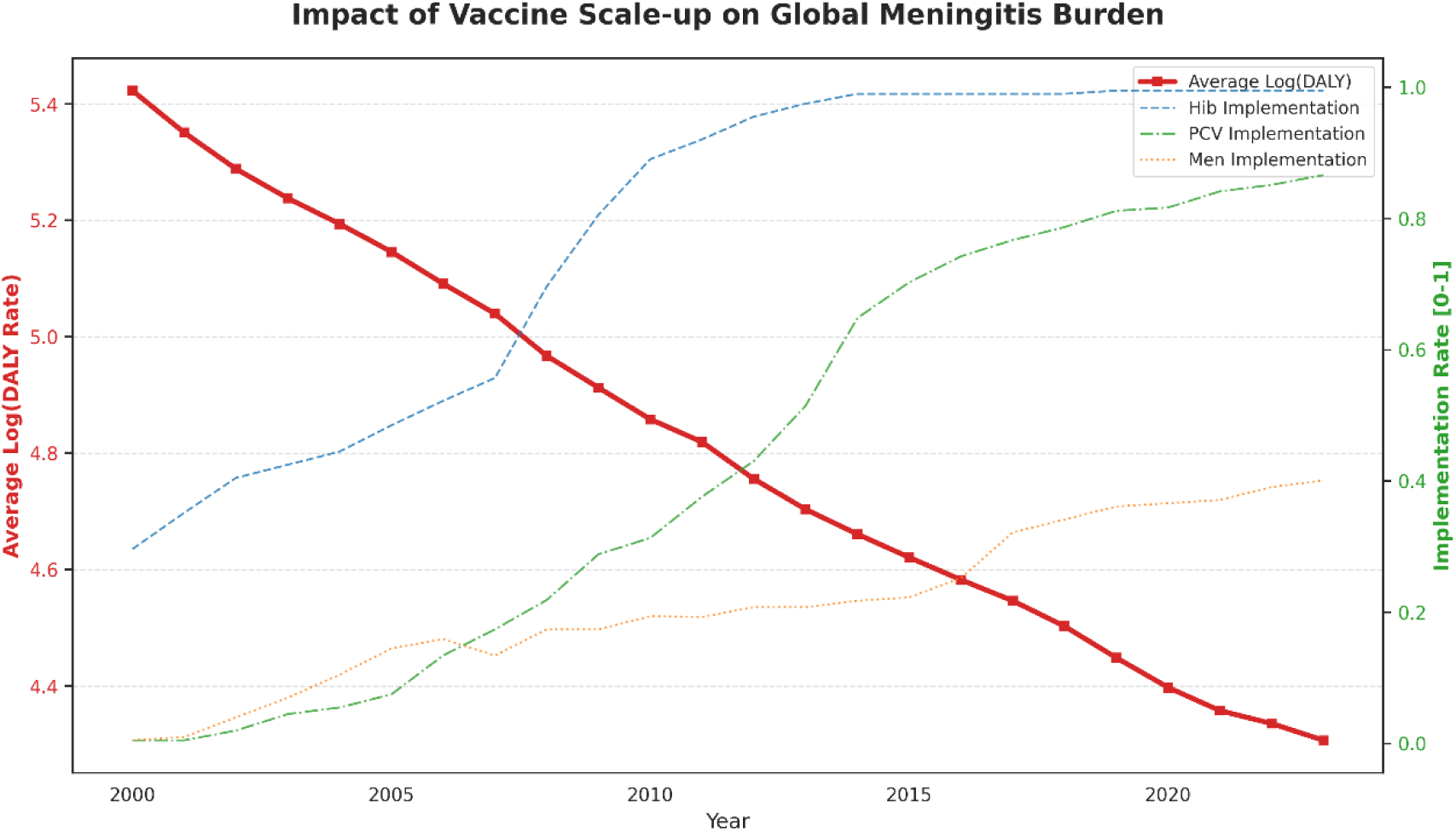
Global trends in meningitis DALY rates and vaccine scale-up, 2000– 2023. Temporal trends in log-transformed meningitis DALY rates across all countries from 2000 to 2023. The solid line represents the mean global DALY trajectory, with shaded areas indicating variability across countries (if applicable). Vertical reference markers (if shown) indicate approximate periods of widespread introduction of Hib, pneumococcal conjugate, and meningococcal vaccines.

**Table 3.** Global association between vaccine introduction and meningitis DALY rates, 2000–2023.

| Variable | Model 1:<br>Baseline | Model 2: Time<br>trend | Model 3: Vaccine-<br>adjusted |
| --- | --- | --- | --- |
| Year (per annum) | — | −0.0497***<br>(−36.16) | — |
| Hib vaccine<br>(post) | — | — | −0.0689*** (−2.72) |
| PCV (post) | — | — | −0.0487** (−2.35) |
| Men vaccine<br>(post) | — | — | 0.0224 (0.84) |
| Observations | 4829 | 4829 | 4829 |
| Within R<sup>2</sup> | 0.000 | 0.779 | 0.328 |
| Fixed effects | Country | Country, year | Country, year |

Data are $\beta$ coefficients (t statistics). \*\*\*p<0.001, \*\*p<0.01, \*p<0.05. Associations between vaccine introduction and log-transformed meningitis DALY rates using two-way fixed effects regression models. Model 2 estimates global temporal trends; Model 3 includes vaccine indicators. All models include country fixed effects; models 2 and 3 additionally include year fixed effects. Standard errors are clustered at the country level.

### 3.3. Impact of Conjugate Vaccines

In models incorporating vaccine indicators (Table 3, Model 3), introduction of the Hib vaccine was associated with a significant reduction in DALY rates (β = −0.0689, *p* < 0.01), corresponding to an estimated 6.7% lower DALY rate in disease burden. Similarly, PCV introduction was associated with a 4.8% reduction in DALY rates (β = −0.0487, *p* < 0.05). The Men vaccine indicator was not significantly associated with DALY reduction in the global model (β = 0.0224, *p* = 0.40). Results were robust to alternative model specifications, including the use of untransformed health metrics. Sensitivity analyses incorporating GDP per capita and HAQI adjustment yielded similar post-introduction patterns, particularly for Hib and PCV vaccines, supporting the robustness of the primary findings to major time-varying socioeconomic and healthcare system factors (Table 5).

### 3.4. Heterogeneity by Baseline Disease Burden

Stratified analyses revealed substantial heterogeneity in vaccine effects (Table 2). In high-burden countries, both Hib and PCV introduction were associated with large, statistically significant reductions in DALY rates (Hib: β = −0.0962, *p* < 0.001; PCV: β = −0.0949, *p* < 0.01), corresponding to 9–10% decreases in disease burden. In low-burden countries, neither vaccine introduction was significantly associated with changes in DALY rates.

### 3.5. Dynamic effects of vaccine introduction (event-study analysis)

To evaluate the temporal dynamics of vaccine impact and assess the validity of the identifying assumptions underlying the two-way fixed effects model, we conducted an event-study analysis of Hib vaccine introduction (Figure 3).

**Figure 3.**
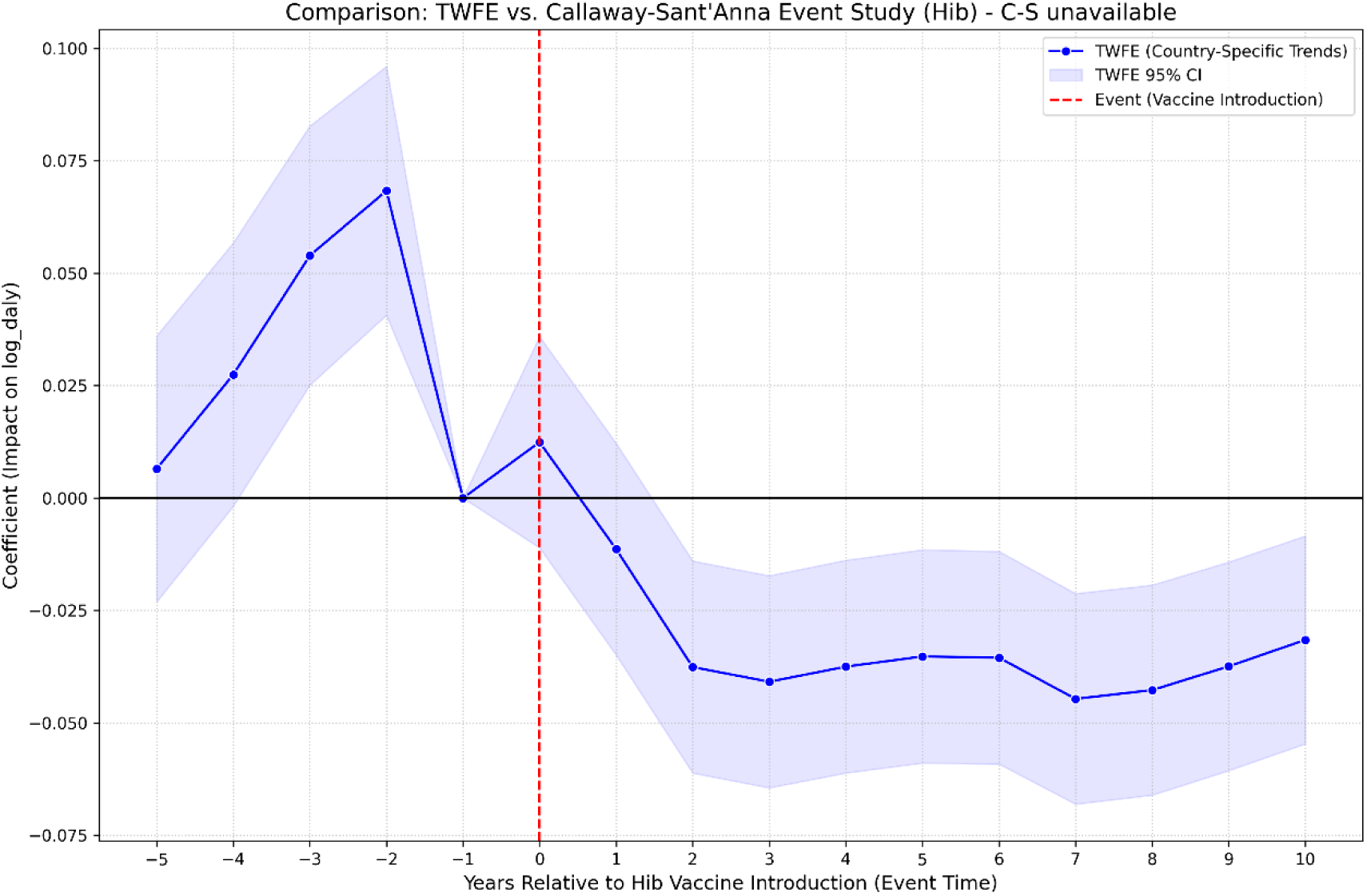
Event-study of Hib vaccine introduction impact on log DALY rates. Coefficients are estimated from an event-study specification incorporating country and year fixed effects with country-specific linear time trends. Event time is defined relative to the year of Hib vaccine introduction, with the year prior to introduction (event time = −1) used as the reference category. Shaded regions represent 95% confidence intervals, and the vertical dashed line indicates vaccine introduction

In the pre-treatment period (event time −5 to −2), we observed a clear upward trend in log DALY rates, with coefficients increasing and reaching approximately 0.07 (p<0.05) two years prior to vaccine introduction. The observed positive pre-treatment trends suggest that vaccine introduction was likely endogenous to worsening meningitis burden trajectories in some settings, limiting strict causal interpretation. Following normalization at the reference period (event time −1), a marked shift in trajectory is observed at the time of vaccine introduction (event time 0), consistent with a structural break in trend. From two years post-introduction onward, the estimated effects become negative and statistically significant (β≈−0.04), and remain relatively stable through event time +10. The corresponding 95% confidence intervals exclude zero from event time +2 onward, indicating a sustained reduction in meningitis burden following vaccine introduction that is unlikely to be explained solely by pre-existing trends.

### 3.6. Regional Analysis: African Meningitis Belt

In the targeted analysis of the 15-country African meningitis belt cohort (Table 4 & Figure 4), PCV introduction was associated with a substantial and statistically significant reduction in DALY rates (β = −0.1833, 95% CI −0.2851 to −0.0815; p < 0.001). This estimate corresponds to an approximate 16.8% decrease in disease burden, representing an effect size nearly four times larger than that observed in the global analysis.

**Figure 4.**
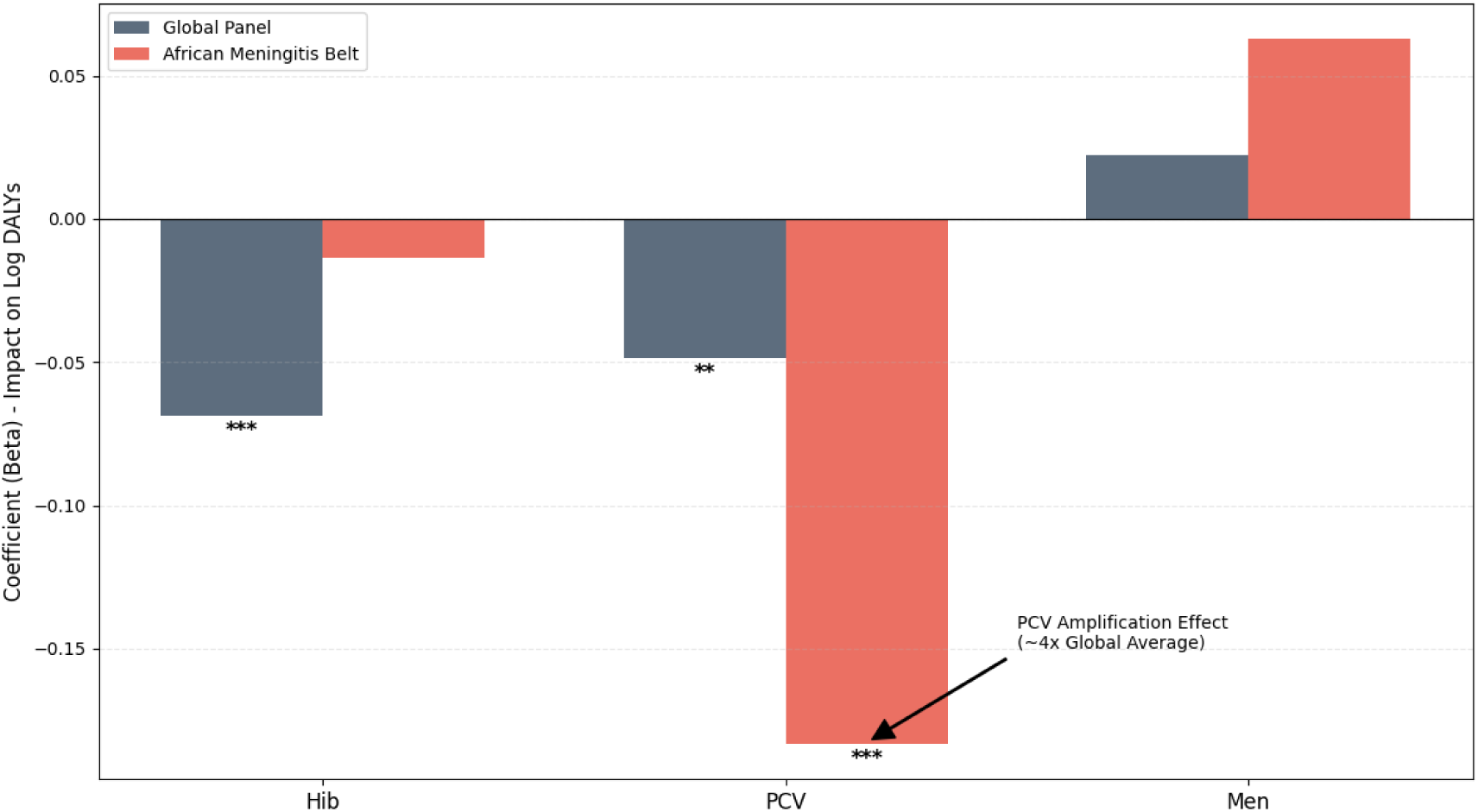
Estimated association between vaccine introduction and meningitis DALY rates globally and in the African meningitis belt. Comparison of regression coefficients for vaccine introduction from two-way fixed effects models. Estimates are shown for global analyses and for the African meningitis belt subgroup. Error bars represent 95% confidence intervals. Negative coefficients indicate reductions in DALY rates following vaccine introduction.

**Table 4.** Association between vaccine introduction and meningitis DALY rates in the African meningitis belt.

| Variable | $\beta$<br>coefficient | Standard<br>error | t<br>statistic | p<br>value | 95% CI |
| --- | --- | --- | --- | --- | --- |
| Hib vaccine<br>(post) | -0.0136 | 0.0665 | -0.20 | 0.838 | -0.1443 to<br>0.1171 |
| **PCV (post)** | **-0.1833\*** | **0.0518** | **-3.54** | **0.0004** | **-0.2851 to**<br>**-0.0815** |
| Men vaccine<br>(post) | 0.0627 | 0.0851 | 0.74 | 0.461 | -0.1044 to<br>0.2299 |

**Note:** $\beta$ coefficients represent the estimated change in log-transformed DALY rates associated with vaccine introduction.
**Model characteristics:** Total observations = 517; Countries = 15; Time periods = 24; Within $R^2 = 0.258$ . All models utilize a two-way fixed effects (TWFE) framework incorporating both country and year fixed effects, with standard errors clustered at the country level. *\* $p < 0.001$ based on $t$ -distribution.*
Association between vaccine introduction and meningitis DALY rates in 15 countries within the African meningitis belt. Estimates are derived from two-way fixed effects models with country and year fixed effects. Standard errors are clustered at the country level.

**Table 5.** Sensitivity-adjusted event-study estimates for vaccine introduction and log DALY rates. . Models adjusted for GDP per capita and Healthcare Access and Quality Index.

| Vaccine | Model | Pre-intro<br>(ET = -2) | Intro<br>(ET = 0) | Early Impact<br>(ET = 2) | Long-term (ET<br>= 5) |
| --- | --- | --- | --- | --- | --- |
| | | $\beta$<br>(95% CI) | $\beta$<br>(95%<br>CI) | $\beta$<br>(95% CI) | $\beta$<br>(95% CI) |
| **Hib** | Adjusted<sup>†</sup> | 0.059*<br>(0.027,<br>0.092) | 0.015<br>(-0.011,<br>0.042) | -0.041 (-0.066,<br>-0.015)* | -0.048 (-0.075,<br>-0.021)* |
| **PCV** | Adjusted<sup>†</sup> | -0.014<br>(-0.041,<br>0.014) | -0.011<br>(-0.039,<br>0.016) | -0.042 (-0.070,<br>-0.014)* | -0.072 (-0.101,<br>-0.043)* |
| **Men** | Adjusted<sup>†</sup> | 0.007<br>(-0.033,<br>0.047) | -0.011<br>(-0.050,<br>0.028) | -0.037 (-0.078,<br>0.005) | -0.044 (-0.090,<br>0.002) |

\*ET: Event Time (Years relative to introduction); CI: Confidence Interval. <sup>†</sup>Adjusted for log GDP per capita and Healthcare Access and Quality Index (HAQI). $p < 0.05$ .

In contrast, no statistically significant associations were identified for Hib (β = −0.0136, p = 0.838) or meningococcal (β = 0.0627, p = 0.461) vaccine introduction within this specific regional model. This suggests that while PCV has had a profound, measurable impact on the meningitis burden in this high-endemicity zone, the annual DALY metrics may not yet capture the episodic or campaign-based impact of meningococcal interventions.

## 4. Discussion

### 4.1. Principal Findings

This longitudinal analysis of global meningitis burden from 2000 to 2023 demonstrates a consistent and statistically significant decline in health loss. Our TWFE models indicate an average annual reduction in log-transformed DALY rates of 4.97%, reflecting substantial progress in global meningitis control.

Because vaccine adoption occurred asynchronously across countries, these estimates should be interpreted as average associations under a staggered-adoption difference-in-differences framework rather than invariant causal treatment effects. This improvement is closely linked to the strategic scale-up of immunization programs, particularly Hib and PCV[2–4]. Sensitivity analyses reveal that the public health impact of these vaccines is concentrated in high-burden countries[5,6]. Notably, substantially larger PCV-associated reductions were observed within the African meningitis belt, where the estimated effect size was nearly four times larger than the global average. The persistence of negative post-introduction coefficients following adjustment for GDP per capita and HAQI suggests that the observed associations are not solely explained by broad improvements in socioeconomic conditions or healthcare access.

### 4.2. Regional Heterogeneity and Vaccine Dynamics

The differential impact of vaccine programs highlights substantial regional heterogeneity. While Hib and PCV demonstrate global significance, Men vaccine indicators did not reach significance in our TWFE models. This discrepancy likely reflects differences in deployment: routine neonatal schedules for Hib and PCV provide continuous coverage, whereas Men vaccines have historically been delivered through campaign-style interventions[14,18]. Annual DALY aggregation may mask the immediate but short-lived reductions associated with episodic Men vaccine campaigns, and the high inter-annual volatility of meningococcal epidemics reduces the signal-to-noise ratio in country-level panel data[16].

In high-burden settings, the introduction of Hib and PCV vaccines each contributed to nearly a 10% reduction in the total burden of meningitis, whereas these effects were not statistically detectable in low-burden settings.

### 4.3. Comparison with Existing Literature

Our findings align with prior evidence demonstrating that routine Hib and PCV immunization programs significantly reduce childhood meningitis burden[2,3]. The pronounced PCV effect in the African belt corroborates earlier epidemiological studies showing the disproportionate impact of pneumococcal disease in this region and the efficacy of mass immunization campaigns in high-risk populations[18–20]. In contrast, Men vaccine rollouts may require sub-national and temporally granular data to detect their full epidemiological impact.

### 4.4. Policy Implications

These findings support prioritisation of vaccine deployment in settings with the highest baseline meningitis burden, where the marginal population-level benefits appear greatest. The substantially larger estimated effects observed in high-burden countries and within the African meningitis belt suggest that targeted immunisation strategies may yield greater reductions in morbidity and premature mortality than uniform allocation approaches.

While the global meningitis burden remains predominantly driven by premature mortality (YLL), the ongoing contribution of long-term disability (YLD) highlights an important epidemiological challenge. As acute mortality declines and survivorship improves, the absolute number of individuals living with post-meningitis neurological sequelae may become more apparent. However, this ecological analysis cannot directly evaluate survivorship trajectories or post-meningitis neurological sequelae. Future longitudinal studies using patient-level outcome data are needed to clarify how reductions in mortality influence long-term disability burden following meningitis.

### 4.5. Strengths and Limitations

The study’s primary strength is its use of a comprehensive, harmonized panel dataset spanning 24 years, enabling robust TWFE estimation. Log-transformations of highly skewed health metrics ensured model validity. Key limitations include:

1. Sample attrition due to exclusion of sub-national aggregates, which may underrepresent localized high-burden zones[4].
2. Residual time-varying confounding remains possible despite sensitivity analyses adjusting for GDP per capita and HAQI, as other unmeasured factors—including sanitation, conflict exposure, healthcare financing, antimicrobial access, and surveillance quality—may also influence meningitis burden trajectories and vaccine adoption timing[12].
3. Temporal resolution, particularly for Men vaccine effects, which may be underestimated due to annual aggregation masking short-term outbreak suppression[16,18].
4. Recent econometric literature has demonstrated that conventional two-way fixed effects estimators may produce biased treatment effect estimates under staggered intervention timing and heterogeneous treatment effects, particularly when already-treated units serve as implicit controls for later-treated units[17,21,22]. Because vaccine introductions occurred asynchronously across countries and the magnitude of vaccine-associated effects likely varied by baseline disease burden and calendar period, the estimated coefficients should be interpreted as average associations rather than definitive causal effects. Although our event-study analyses partially address dynamic treatment timing, future work using modern staggered-adoption estimators, such as the Sun-Abraham or Callaway-Sant’Anna frameworks, would strengthen causal interpretation.

## Data Availability

All data produced in the present study are available upon reasonable request to the authors

## Ethics Committee Approval

This study utilized publicly available, de-identified secondary data from IHME and WHO; thus, it was deemed exempt from institutional review board approval.

## Data Sharing Statement

All health outcome data are available via the GBD Results Tool (http://ghdx.healthdata.org/gbd-results-tool). Vaccine implementation data are available via the WHO Immunization Dashboard.

## Acknowledgments

The author would like to express gratitude to ReseMeds Academy for providing the technical framework, statistical consulting, and research coordination that facilitated this Global study.

## Authorship Statement

The author attests they meet the ICMJE criteria for authorship. Khalid Mohammed Al-Dhayani: Conceptualization, Methodology, Software, Data curation, Formal analysis, Investigation, Writing - original draft, Writing - review & editing, Visualization, Project administration.

## Supplementary

**Supplementary Figure S1.**
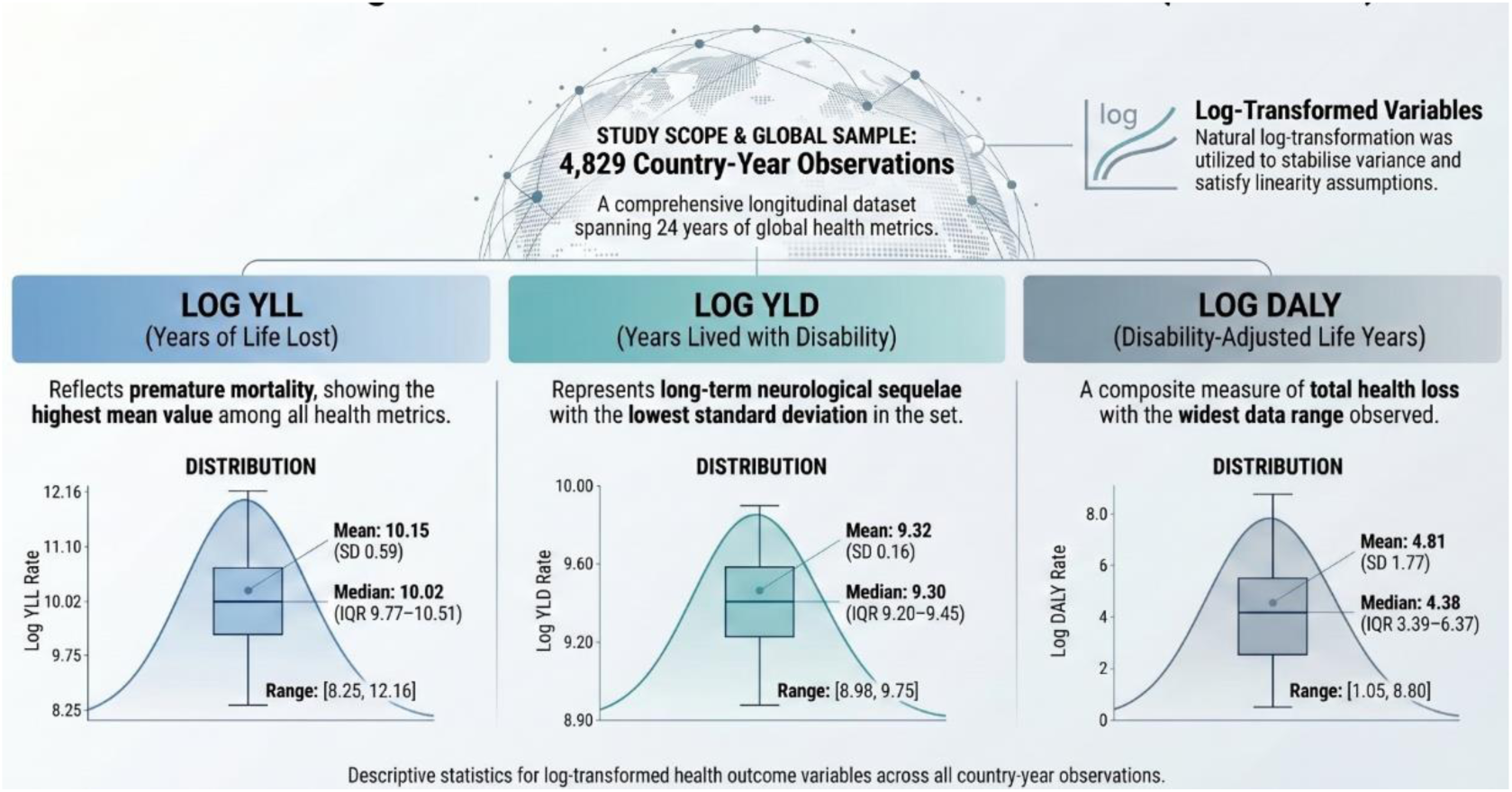
Distribution of log-transformed global meningitis health outcomes (2000–2023). The figure displays the density and box plot distributions of the natural log-transformed metrics across 4,829 country-year observations: Years of Life Lost (YLL), Years Lived with Disability (YLD), and Disability-Adjusted Life Years (DALYs). Natural log-transformation was applied to the raw, right-skewed global health metrics to stabilize variance and satisfy the linearity and homoscedasticity assumptions required for the two-way fixed effects parametric modeling. Descriptive statistics, including mean, standard deviation (SD), median, and interquartile range (IQR), are overlaid for each transformed outcome.

**Supplementary Figure S2.**
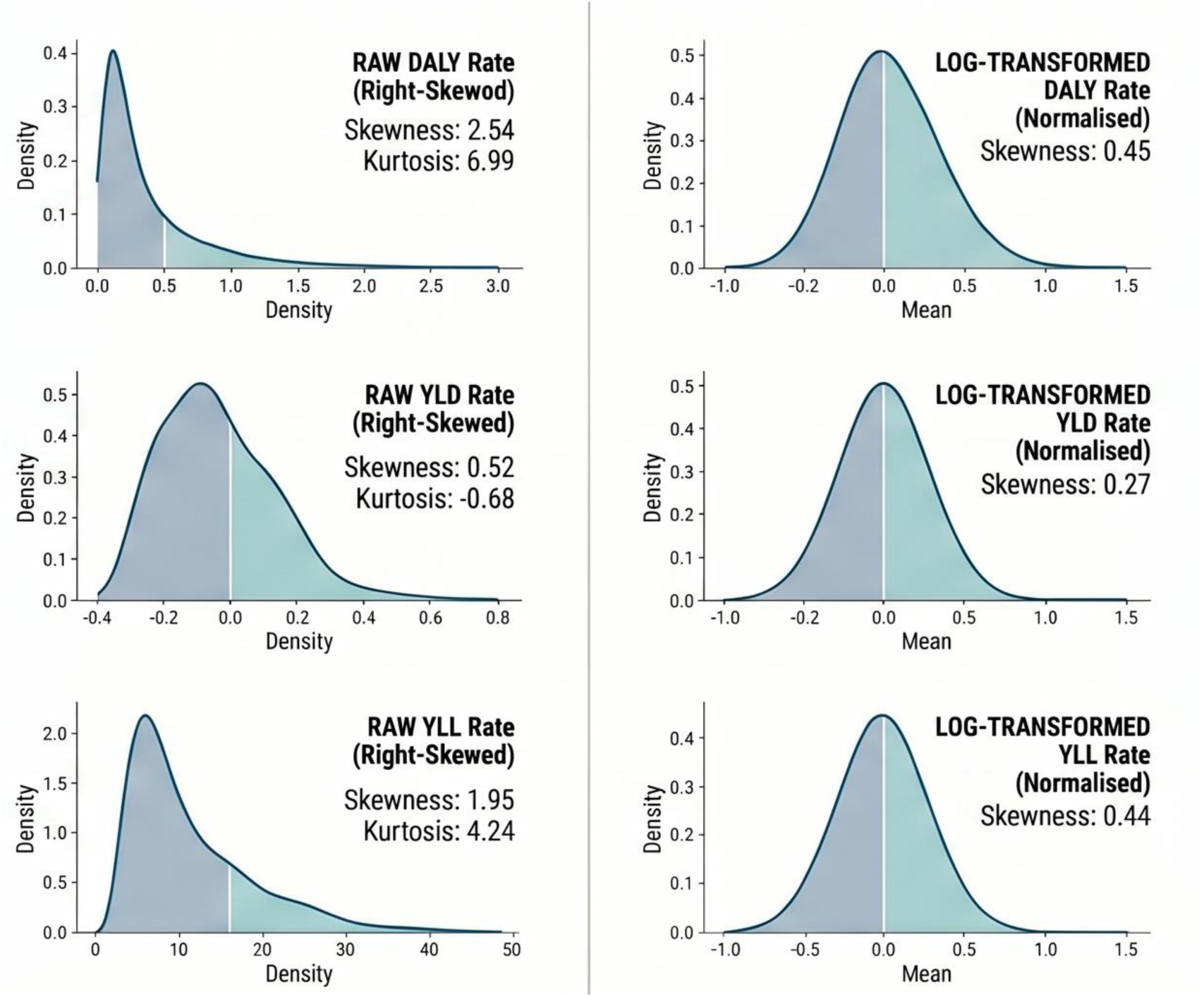
Distributional properties of meningitis health metrics before and after natural logarithmic transformation (2000–2023). Density plots comparing the distributions of raw health metrics (left column) against their natural log-transformed counterparts (right column) across 4,829 country-year observations. Raw Disability-Adjusted Life Year (DALY) and Years of Life Lost (YLL) rates exhibited severe right-skewness and high kurtosis, violating the normality and homoscedasticity assumptions required for parametric panel regression. Natural log-transformation successfully stabilized the variance, reducing the skewness of DALY rates from 2.54 to 0.45, resulting in approximately normal distributions suitable for two-way fixed effects (TWFE) econometric modeling.

